# Use of Renal Duplex To Detect Renal Artery Stenosis in Fibromuscular Dysplasia

**DOI:** 10.64898/2026.09.16.26363272

**Authors:** Fahad Alkhalfan, Meghann McCarthy, Alliefair Scalise, Hannah Abroe, Anu Aggarwal, Edelyn (Huijin) Park, Pulkit Chaudhury, Scott J. Cameron, Deborah Hornacek, Christopher Bajzer, Natalia Fendrikova Mahlay

## Abstract

**Introduction:** Duplex ultrasonography is widely used to evaluate renal artery stenosis (RAS), but diagnostic criteria are largely derived from atherosclerotic RAS and may not apply to fibromuscular dysplasia (FMD). We evaluated associations between renal duplex parameters and hemodynamically significant RAS in patients with FMD.

**Methods:** Patients with renal FMD who underwent renal duplex followed by renal artery angiography within 3 months were included. Duplex parameters were compared between arteries with and without RAS (Pd/Pa<0.9, fractional flow reserve (FFR)<0.8, systolic pressure gradient >20 mm Hg, or %stenosis >70%). Receiver operating characteristic (ROC) analysis was used to evaluate diagnostic performance. Changes following angioplasty were compared.

**Results:** 55 renal arteries from 33 patients with renal FMD were included. Peak systolic velocity (PSV), end diastolic velocity (EDV), and acceleration time (AT) were higher in arteries with RAS. Resistive index (RI) was lower in RAS. Renal aortic ratio (RAR) did not differ between groups. PSV demonstrated good discrimination with a cut-off of 275 cm/sec yielding 79% sensitivity, 78% specificity, and an area under the curve (AUC) of 0.81. Incorporating additional parameters did not improve performance compared with PSV alone. Following angioplasty, PSV, EDV, and AT decreased, RI increased, RAR remained unchanged.

**Conclusion:** In renal FMD, duplex ultrasound parameters, particularly PSV and RI, are associated with hemodynamically significant RAS. However, overlap in PSV between arteries with and without RAS suggests that PSV alone may not reliably determine lesion significance. These findings support duplex ultrasonography for evaluating renal FMD while highlighting the need for FMD-specific diagnostic criteria.

## Introduction

Hypertension, characterized by a persistent elevation in arterial blood pressure, is the leading preventable cause of premature death worldwide.(1) Primary (essential) hypertension represents 85-95% of human cases and does not have an identified singular etiology.(2) Renovascular hypertension is one of the most common causes of secondary hypertension and often results from renal artery stenosis (RAS).(3) Although atherosclerosis is the predominant cause of RAS, nonatherosclerotic conditions, including fibromuscular dysplasia (FMD), account for many cases.(4) Fibromuscular dysplasia is believed to be a nonatherosclerotic, noninflammatory arterial disease with a female preponderance that can result in arterial stenosis, occlusion, aneurysm, or dissection.(4) FMD affects the renal arteries in 86% and 91% of cases in the United States Registry and EU/International Registry for Fibromuscular Dysplasia, respectively.(5) Hypertension in patients with FMD may be diagnosed at an age that overlaps with that of patients with essential hypertension.(6,7) Renovascular hypertension as a consequence of FMD should be suspected in patients, particularly women, with early onset, resistant, accelerated, or malignant hypertension and in those with known FMD in another vascular territory.(8) While computed tomography angiography (CTA) is commonly used as the initial imaging modality for the diagnosis of renal FMD, invasive catheter-based angiography remains the gold standard, particularly when endovascular intervention is being considered.

Duplex ultrasonography is widely used as the initial imaging modality for patients with suspected renovascular hypertension and offers many advantages including its noninvasive nature, reduced cos, and high-quality physiological imaging without exposing patients to ionizing radiation or iodinated contrast agents. (9,10) In patients with atherosclerotic RAS, renal duplex ultrasonography criteria, including a peak systolic velocity (PSV) of > 200 cm/sec and a renal-aortic ratio (RAR) of ≥ 3.5 have demonstrated high diagnostic accuracy for significant RAS (≥ 60%). Reported sensitivities and specificities range from 95-97% and 79-90%, respectively, for PSV, 91-92% and 62-91%, respectively, for RAR.(11,12) However, the diagnostic criteria currently used for renal artery stenosis were largely developed and validated in patients with atherosclerotic RAS, in whom the lesions typically involve the ostium or proximal renal artery.(13) In contrast, FMD most commonly affects the mid to distal renal artery and often produces multifocal lesions with a characteristic “string-of-beads” appearance appreciable on angiography and sometimes color Doppler imaging, as well as turbulence and elevated velocities at the mid to distal arterial segments on ultrasound.(6) Consequently, the duplex ultrasound criteria used to diagnose atherosclerotic RAS are not applicable to renal artery stenosis caused by FMD due to dissimilarities between these arterial pathologies. Utilizing a cohort of patients with renal FMD seen in our healthcare system, we aimed to determine the utility of ultrasound in identifying hemodynamically significant renal artery stenosis due to FMD.

## Methods

### Patient Selection

This retrospective study included patients with renal FMD seen at the Cleveland Clinic FMD clinic that evaluates > 2000 patients annually with FMD, allowing for the study of patients that are well phenotyped. This study identified all patients with renal FMD who had both a renal angiogram and a renal duplex ultrasound within the three-month period prior to their angiogram. All patients meeting these criteria between 2004 and 2023 were included. Arteries with prior stenting or surgical bypass were excluded from the analysis. All renal duplex ultrasound examinations were performed in the same Intersocietal Accreditation Commission (IAC)-accredited vascular laboratory.

### IRB Statement

This study was conducted in accordance with the ethical principles of the Declaration of Helsinki and approved by the Cleveland Clinic Institutional Review Board.

### Identification of Hemodynamically Significant RAS On Catheter Angiography

We limited our analysis to vessels with renal FMD confirmed on renal angiogram. If a patient had two vessels with FMD, each affected vessel was analyzed separately. We defined renal FMD stenosis as being hemodynamically significant if it met at least one of the following criteria: (1) Systolic pressure gradient greater than or equal to 20 mm Hg across a stenotic lesion; (2) Fractional flow reserve (FFR) ≤ 0.80; (3) Distal pressure to aortic pressure ratio (Pd/Pa ratio) < 0.9; (4) Stenosis ≥ 70% as determined by the operator.(8,14) All data were extracted from angiographic reports, which included at least one quantitative parameter per vessel, with some reports providing multiple measures.

### Performance of US Parameters in Identifying RAS

The vascular laboratory where each study was conducted is a high volume (>70,000 studies annually), Intersocietal Commission for the Accreditation of Vascular Laboratories (ICAVL)-accredited laboratory. Images were captured by professional and registered vascular technologists (RVT) not involved in this study. Image interpretation was by multiple physicians not involved in this study who hold the Registered Physician in Vascular Interrelation (RPVI) credential.

After identifying the peak systolic velocity (PSV), end diastolic velocity (EDV), acceleration time (AT), resistive index (RI), and RAR, we constructed univariable logistic regression models to assess the association between each of the four parameters and the presence of RAS. Each parameter was assessed separately. Because some patients contributed more than one renal artery to the analysis, observations were not assumed to be independent, and all regression models were fit with standard errors clustered at the patient level. For each parameter, Receiver Operating Characteristic (ROC) curves were generated and the area under the curve (AUC) was calculated. Optimal cut-off values were selected from ROC analyses by identifying the threshold with the best combined sensitivity and specificity (Youden index). The STATA program “cutpt” was used to calculate the Youden index. Model calibration was assessed using the Hosmer-Lemeshow goodness-of-fit test.

### Building the Prediction Model

We built two different models: (1) Model 1 only included parameters with an AUC greater than 0.7; (2) Model 2 only included PSV. PSV was selected for Model 2 because it is the primary parameter used to identify hemodynamically significant RAS in atherosclerotic disease. Model discrimination was assessed by calculating the AUC. Additionally, Akaike Information Criterion (AIC) and Bayesian Information Criterion (BIC) were calculated to measure model quality, with a lower value indicating a better model. To further evaluate the performance and generalizability of our model, we employed five-fold cross-validation using the “cvauroc” command in Stata, specifying clustering at the patient level. The dataset was randomly divided into five equal-sized groups. In each iteration, one group was used as the validation set while the other four were used for training. This process was repeated five times to ensure that each group was used for validation only once. The results from each iteration were averaged to obtain an internally validated estimate of model discrimination. Bootstrap bias-corrected 95% confidence interval were calculated for the cross-validated AUC.(15) Finally, the Wald test was used to assess whether the addition of variables beyond PSV added additional predictive value to the model (Model 1 vs Model 2).

### Post Intervention Imaging

We evaluated changes in ultrasound parameters in arteries with hemodynamically significant renal artery stenosis following intervention. For each treated artery, the first ultrasound performed within 1-year post-intervention was identified. Paired t-tests were used to compare ultrasound parameters before and after intervention.

### Sensitivity Analysis

Due to limitations of identifying hemodynamically significant stenosis in patients with FMD using degree of stenosis alone, an additional sensitivity analysis was conducted in arteries that had a physiological measurement (FFR, systolic pressure gradient or Pd/Pa). The performance of each ultrasound measure separately as well as models 1 and 2 were evaluated in this subset of arteries. Additionally, we compared each parameter as well as both models in arteries without prior angioplasty.

The analysis was done using STATA/MP Version 16 (StatCorp, College Station, Tx). Continuous variables were compared using the Student’s *t*-test or Wilcoxon rank-sum test, as appropriate, and categorical variables were compared using the chi-square test or Fisher’s exact test. A two-sided p-value of < 0.05 was considered statistically significant.

## Results

### Patient Characteristics

A total of 55 renal arteries with FMD from 33 patients were identified and included in the study. The mean age was 47.7 years, all patients were female, and all had a history of hypertension (Table 1). Hemodynamically significant RAS was present in 14 of 55 renal arteries (25.5%) representing 12 patients, including 2 patients with bilateral disease. Fifty arteries (82%) underwent physiologic assessment (Pd/Pa, FFR, or systolic pressure gradient) and were included in the additional sensitivity analysis. Patients with hemodynamically significant stenosis were younger, whereas all other baseline characteristics were similar between groups (**Table 2)**.

**Table 1.** Baseline Characteristics of Patients with Renal FMD.

| Characteristic | Overall (N=33) |
| --- | --- |
| Age (years), mean ± SD | 47.7 ± 15.8 |
| Female sex, n (%) | 33 (100) |
| BMI, mean ± SD | 23.11 ± 4.67 |
| History of hypertension, n (%) | 33 (100) |
| Prior angioplasty, n (%) | 47.7 (15.8) |
| Medications at time of enrollment in the registry, n (%) |  |
| ACEi/ARB | 20 (61) |
| Beta blocker | 16 (48) |
| Diuretic | 9 (27) |
| Alpha 2 antagonist | 2 (6) |
| Aspirin | 22 (67) |
| Clopidogrel | 6 (18) |
| Statin | 7 (21) |

**Table 2.** Baseline characteristics by artery type (stenosis vs no stenosis)*

| Characteristic | No stenosis (N = 41) | Stenosis (N = 14) | P-value |
| --- | --- | --- | --- |
| Age (Years), mean ± SD | 54.0 (12.2) | 35.2 (13.2) | < 0.001 |
| Female, n (%) | 41 (100) | 14 (100) | 1 |
| BMI, mean ± SD | 23.4 (5.5) | 22.1 (1.6) | 0.46 |
| History of hypertension, n (%) | 41 (100) | 14 (100) | 1 |
| Prior angioplasty, n (%) | 9 (22%) | 3 (21%) | 0.97 |
| Medications at time of enrollment in the registry, n(%) |  |  |  |
| ACEi/ARB | 25 (61) | 6 (43) | 0.59 |
| Beta blocker | 21 (51) | 2 (14) | 0.16 |
| Diuretic | 14 (34) | 2 (14) | 0.16 |
| Alpha 2 antagonist | 2 (5) | 0 (0) | 0.4 |
| Aspirin | 32 (78) | 9 (64) | 0.31 |
| Clopidogrel | 8 (20) | 3 (21) | 0.88 |
| Statin | 12 (29) | 0 (0) | 0.022 |

\* Patients with bilateral renal artery FMD contributed two arteries to the analysis.

### Ultrasound Parameters

#### Peak Systolic Velocity

The mean peak systolic velocity was 257 cm/s (± 95.3 cm/s). Compared with vessels without hemodynamically significant RAS, vessels with RAS had a higher mean PSV (RAS vs no RAS: 332.8 ± 108.0 cm/s vs 231.3 ± 75.9 cm/s) (Figure 1A). On logistic regression analysis, each 1 cm/s increase in PSV was associated with a higher odds of hemodynamically significant RAS (OR 1.014, 95% confidence interval (CI) 1.003 – 1.025), p = 0.016). ROC analysis yielded an AUC of 0.81 (Figure 2A). The optimal PSV threshold identified by the Youden index was 275.5 cm/s, corresponding to a sensitivity of 79% and specificity of 78%. Model calibration was acceptable, with no evidence of poor fit (Hosmer-Lemeshow goodness-of-fit test p value = 0.11). Notably, the three arteries classified as not having hemodynamically significant RAS despite a PSV greater than 350 cm/s had systolic pressure gradients of 14, 16, and 18 mm Hg.

**Figure 1.**
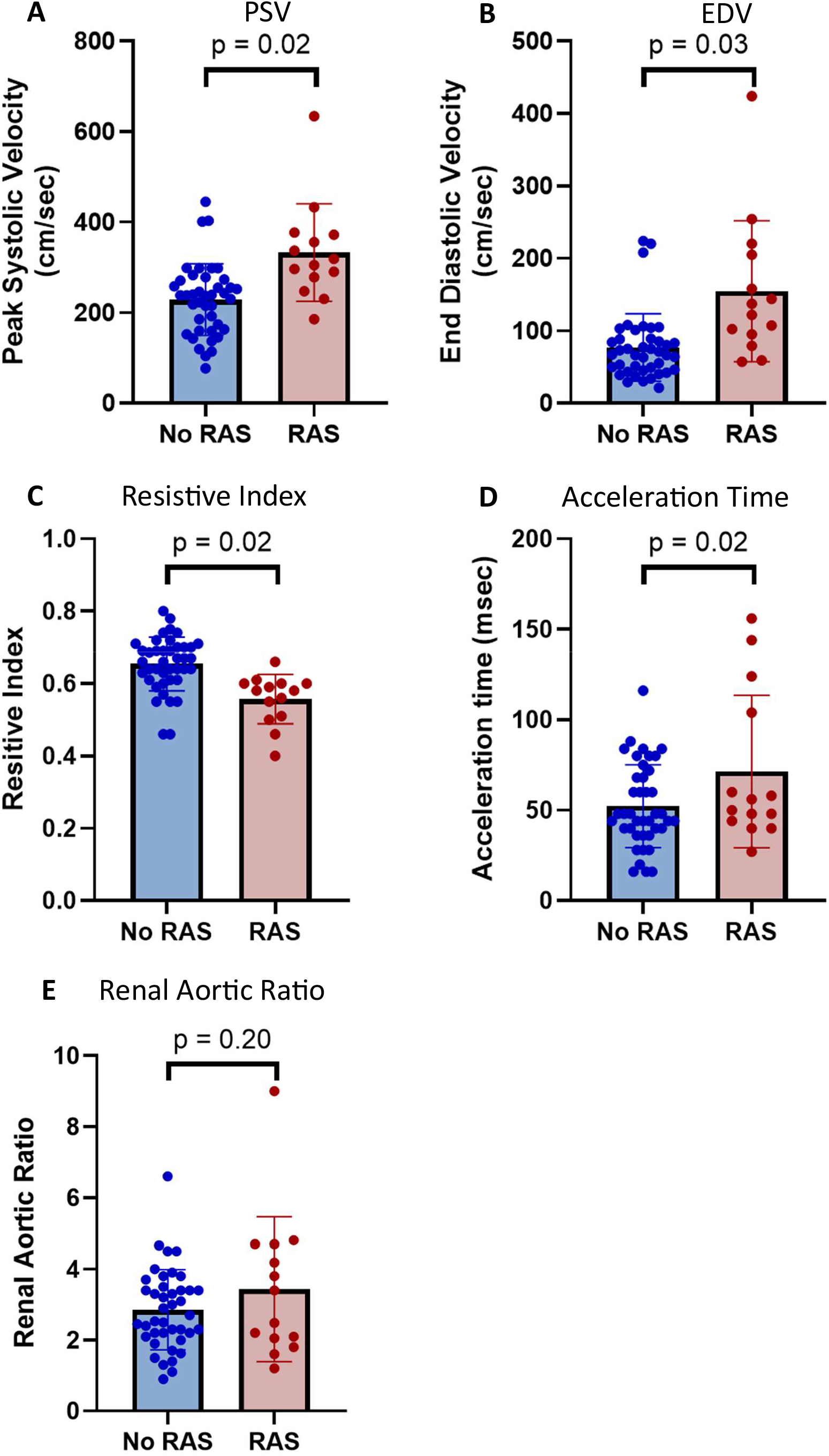
Ultrasound Parameters by Stenosis Status

**Figure 2.**
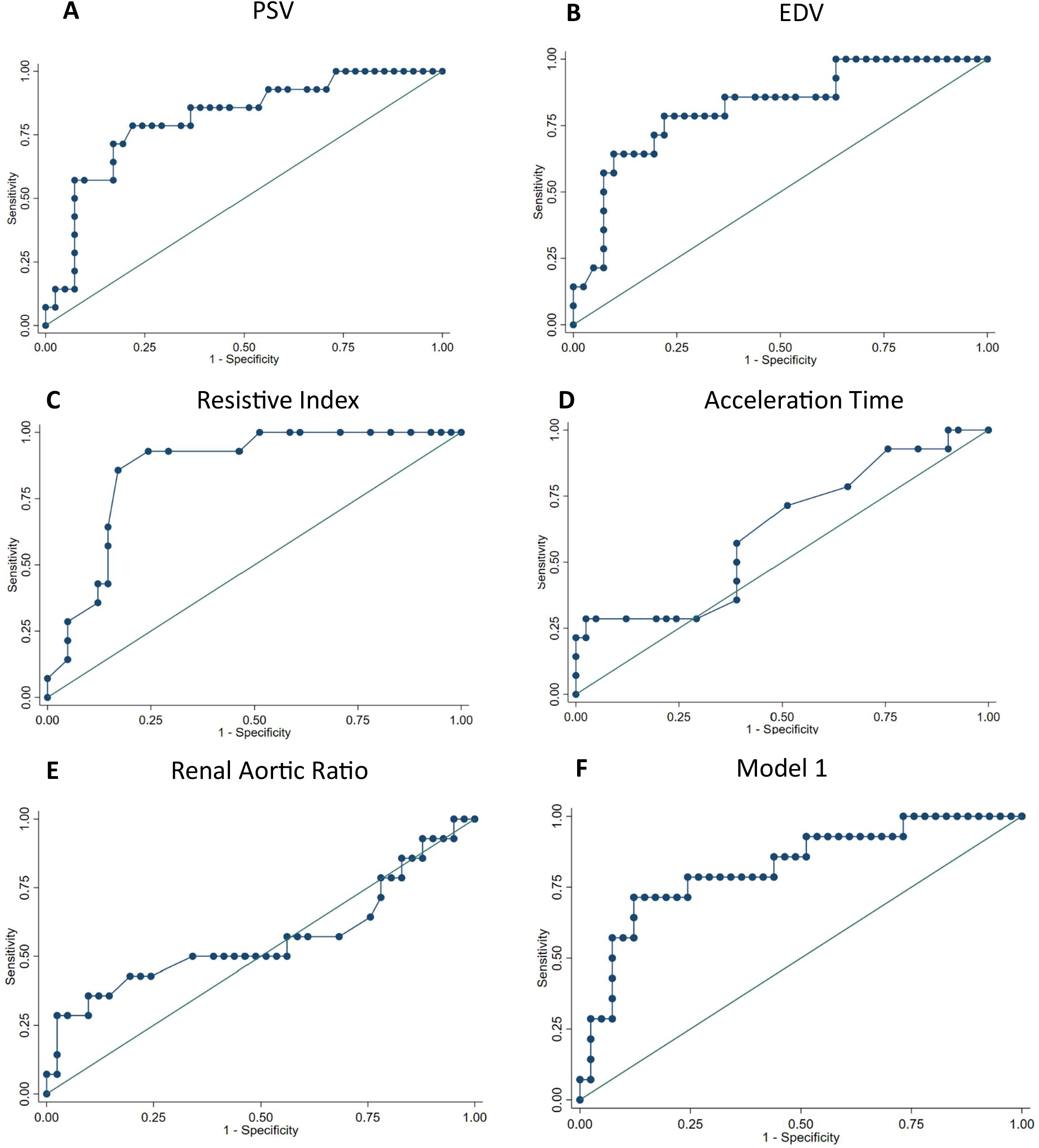
ROC Curves for Each Ultrasound Parameter and Model 1 (PSV, EDV, Resistive Index)

#### End Diastolic Velocity

The mean end diastolic velocity was 97.2 ± 70.7 cm/s. Compared with vessels without hemodynamically significant RAS, vessels with RAS had higher EDV values (RAS vs no RAS: 154.5 ± 97.6 cm/s vs 77.7 ± 45.9 cm/s) (Figure 1B). On logistic regression analysis, each 1 cm/s increase in EDV was associated with an increased odds of hemodynamically significant RAS (OR 1.02, 95% CI 1.002 – 1.034 p = 0.03, p = 0.03). ROC curve analysis yielded an AUC of 0.82 (Figure 2B). The optimal EDV threshold identified by the Youden index was 92 cm/s, corresponding to a sensitivity of 79% and specificity of 78%. Model calibration was acceptable, with no evidence of poor fit on the Hosemer-Lemeshow goodness-of-fit test (p = 0.24).

#### Resistive Index

The mean RI was 0.63 (± 0.08). Compared with vessels without hemodynamically significant RAS, vessels with RAS had lower RI (RAS vs no RAS: 0.56 ± 0.07 vs 0.65 ± 0.07) (Figure 1C). On logistic regression analysis, each 0.01 increase in RI was associated with a decreased odds of hemodynamically significant RAS (OR 0.84, 95% CI 0.72 – 0.98, p = 0.02). ROC analysis yielded an AUC of 0.86 (Figure 2C). The optimal RI threshold by the Youden index of 0.605 corresponded to a sensitivity of 86% and specificity of 83%. There was no evidence of poor fit on the Hosmer-Lemeshow goodness-of-fit test (p = 0.19).

#### Acceleration Time

The mean AT was 57.1 milliseconds (± 29.8 milliseconds). Compared with vessels without hemodynamically significant RAS, vessels with RAS had higher AT (RAS vs no RAS: 71.4 ± 42.1 milliseconds vs 52.3 ± 22.9) (Figure 1D). On logistic regression analysis, each 1 millisecond increase in AT was associated with increased odds of hemodynamically significant RAS (OR 1.02, 95% CI 1.002 – 1.04, p = 0.02). ROC analysis yielded an AUC of 0.62 (Figure 2D). The optimal AT threshold by the Youden index of 96 milliseconds corresponded to a sensitivity of 29% and specificity of 98%. There was no evidence of poor fit on the Hosmer-Lemeshow goodness-of-fit test (p = 0.051).

#### Renal Aortic Ratio (RAR)

The mean RAR was 2.7 (± 1.42). Compared with vessels without hemodynamically significant RAS, vessels with RAS had similar RAR (RAS vs No RAS: 2.94 ± 2.0 vs 2.7 ± 1.13) (Figure 1E). On logistic regression analysis, the association between RAR and RAS was not significant (OR 1.31, 95% CI 0.86 – 2.00, p = 0.20). ROC analysis yielded an AUC of 0.56 (Figure 2E). The optimal RAR threshold identified by the Youden index of 4.6 corresponded to a sensitivity of 29% and specificity of 98%. There was no evidence of poor fit on the Hosmer-Lemeshow goodness-of-fit test (p = 0.29).

### Evaluation of Prediction Models

Model 1 consisted of the three variables that had an AUC of 0.7 or greater (PSV, EDV, resistive index). Model 2 consisted of only PSV. Both models performed similarly, however, Model 2 had slightly lower AIC and BIC (**Table 3, Figure 1A and 1F**). Additionally, the Wald test was not significant (p = 0.23), indicating that adding both EDV and resistive index did not provide incremental value beyond PSV alone.

**Table 3.**
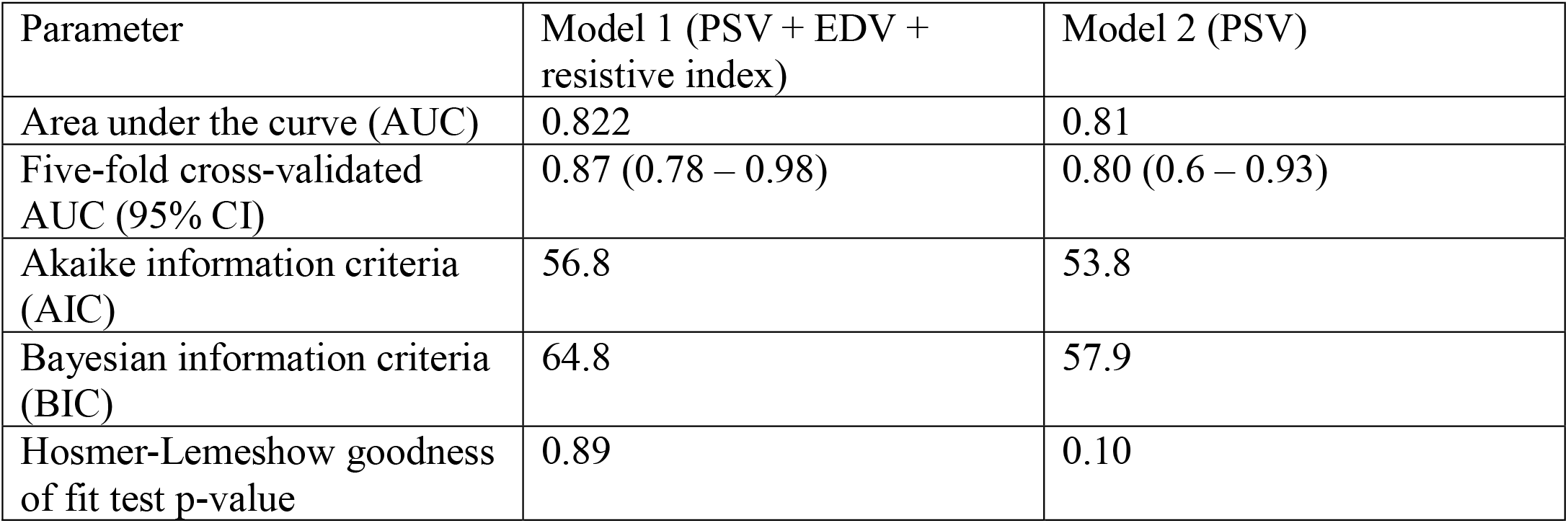
Evaluation of Predictive Capabilities of Models 1 and 2.

### Post Intervention Imaging

Of the 14 arteries with hemodynamically significant RAS, 12 underwent balloon angioplasty and had follow up duplex ultrasound within one year of the procedure (Median: 45 days, interquartile range: 29 – 276 days). As shown in **Figure 3**, post-intervention ultrasound demonstrated a decrease in PSV, EDV, and acceleration time, along with an increase in resistive index. There was no significant difference in RAR before and after intervention.

**Figure 3.**
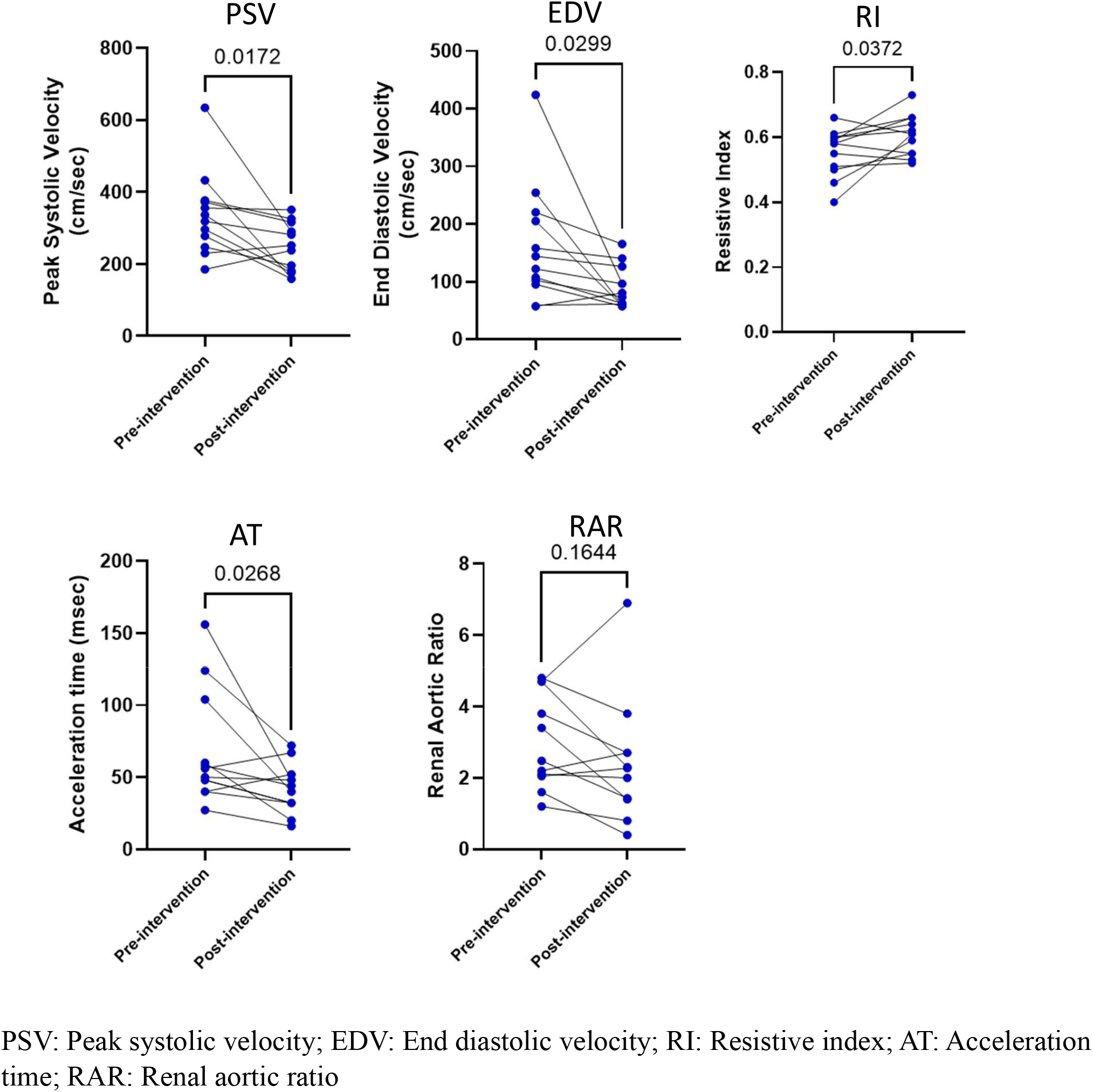
Change in ultrasound parameters after treatment of hemodynamically significant renal FMD

### Sensitivity Analysis

#### Physiologic Assessment

Of the 55 arteries, 50 had at least one hemodynamic measure. As shown in **Supplemental Table 1**, PSV, EDV, and resistive index demonstrated similar performance, whereas acceleration time was not significantly associated with hemodynamically significant RAS. The predictive models showed comparable performance, with slightly lower AIC and BIC as compared to the model built on the entire dataset. Importantly, adding EDV and resistive index did not incrementally improve predictive performance beyond PSV alone (**Supplemental Table 2**).

#### Arteries Without Prior Angioplasty

Forty three out of the 55 arteries in this cohort did not undergo angioplasty previously. As shown in Supplemental Table 3, with the exception of EDV, the rest of the associations that were observed in the main cohort remained statistically significant (PSV, RI, AT). Additionally, Models 1 and 2 performed similarly to what was observed in the main cohort, although the five-fold cross validation yielded an AUC of 0.78 (Supplemental Table 4).

## Discussion

In this study of patients with renal FMD, several renal duplex parameters were associated with hemodynamically significant RAS confirmed by invasive angiography. Specifically, PSV, EDV, resistive index, and acceleration time were significantly associated with the presence of hemodynamically significant RAS with all but AT demonstrating an AUC > 0.7. A predictive model using PSV alone performed comparably to a model using PSV, EDV, and resistive index for identifying arteries with hemodynamically significant RAS. Importantly, in arteries with hemodynamically significant RAS that underwent balloon angioplasty and had follow up imaging, revascularization was associated with reduction in PSV, EDV and AT, accompanied by an increase in RI .

In our cohort, a PSV cut-off of 275.5 cm/s demonstrated a sensitivity of 79% and specificity of 78% in identifying hemodynamically significant RAS. Interestingly, PSV does not appear to be the sole predictor of hemodynamically significant RAS, as there were arteries with PSV values greater than 350 cm/s without RAS, whereas others had PSV less than 250 cm/s despite the presence of RAS. Furthermore, although PSV decreased following angioplasty in arteries with RAS, some post-procedure velocities remained above 300 cm/s. Unlike in atherosclerotic RAS, RAR was not significantly associated with RAS in our cohort. Collectively, these findings suggest that although PSV may be useful in identifying hemodynamically significant RAS in patients with FMD, elevated PSV alone may not reliably identify arteries with hemodynamically significant disease.

Using what is, to our knowledge, the largest reported cohort of patients with FMD evaluating renal artery duplex sonography against invasive angiographic assessment in patients with fibromuscular dysplasia, we demonstrate significant differences in duplex ultrasound parameters between hemodynamically significant and non-significant renal artery stenoses. Unlike atherosclerotic renal artery disease, the role of duplex ultrasonography in identifying significant stenosis in FMD remains incompletely defined. While renal duplex ultrasonography should not be used as the sole determinant for proceeding to renal angiography when otherwise clinically indicated, our findings suggest that it may provide valuable pre-procedural information regarding the likelihood of identifying hemodynamically significant stenosis. Furthermore, the observed reductions in PSV and EDV following angioplasty suggest that serial duplex ultrasonography may have a role in post-procedural surveillance. Specifically, a progressive increase in PSV on follow-up imaging could potentially indicate restenosis, although this hypothesis cannot be confirmed with the current dataset. Prospective studies are needed to validate these findings and further define the role of duplex ultrasonography in the diagnosis and longitudinal surveillance of renal artery stenosis in patients with FMD.

While this study does have many strengths, including a relatively large population of FMD patients, and the use of a single high-volume IAC-accredited vascular laboratory with extensive expertise in evaluation of fibromuscular dysplasia, it also comes with limitations. First, it is a retrospective observational study of patients with fibromuscular dysplasia who underwent duplex ultrasonography within three months prior to renal angiography. This inclusion criteria may have introduced selection bias, as duplex ultrasonography is not routinely performed immediately before angiography in clinical practice. Second, this was a single-center study, and our findings have not been externally validated in an independent cohort. Third, although hemodynamic assessment was available for most arteries, 5 of the 55 arteries did not have a documented hemodynamic measurement. Reassuringly, sensitivity analyses yielded similar results. Additionally, we intentionally included arteries with prior angioplasty as such history would not have affected the decision to undergo renal artery angiography when otherwise medically indicated. However, the sensitivity analysis demonstrates that the majority of findings, including the association between PSV and RAS, remained significant when only looking at arteries without prior angioplasty. Although the hemodynamic parameters used to define lesion significance were concordant in arteries with more than one assessment available, reliance on multiple metrics rather than a single standardized physiologic reference introduces heterogeneity and represents an additional study limitation. Finally, the arteries included in this study are derived from patients who underwent renal artery angiography and therefore may not be representative of the broader population of patients with renal artery FMD.

## Conclusion

In a cohort of patients with renal artery FMD who had both renal duplex ultrasound and renal artery angiography, a cut-off PSV of 275 cm/s demonstrated 79% sensitivity and 78% specificity for identifying hemodynamically significant RAS. Further studies are required to validate the role of duplex ultrasonography in identifying hemodynamically significant RAS in patients with FMD.

## Data Availability

The data underlying this study are not publicly available because they contain protected health information and could compromise patient privacy. Access to these data is restricted in accordance with institutional and patient privacy requirements.

